# Biomonitoring of perfluoroalkyl and polyfluoroalkyl substances (PFAS) from the Survey of the Health of Wisconsin (SHOW) 2014-2016 and comparison with the National Health and Nutrition Examination Survey (NHANES)

**DOI:** 10.1101/2023.02.14.23285850

**Authors:** Amy Schultz, Noel Stanton, Rachel Pomazal, Meshel Lange, Roy Irving, Jonathan Meiman, Brandon Shelton, Kristen CM Malecki

## Abstract

**Background:** Per- and polyfluoroalkyl substances (PFAS) are a growing class of manufactured chemical compounds found in a variety of consumer products. PFAS have become ubiquitous in the environment and were found in many humans sampled in the United States (U.S.). Yet, significant gaps in understanding statewide level exposures to PFAS remain.

**Objective:** The goals of this study are to establish a baseline of exposure at the state level by measuring PFAS serum levels among a representative sample of Wisconsin residents and compare to United States National Health and Nutrition Examination Survey (NHANES).

**Methods:** The study sample included 605 adults (18+ years of age) selected from the 2014-2016 sample of the Survey of the Health of Wisconsin (SHOW). Thirty-eight PFAS serum concentrations were measured using high-pressure liquid chromatography coupled with tandem mass spectrometric detection (HPLC-MS/MS) and geometric means presented. Weighted geometric mean serum values of eight PFAS analytes from SHOW were compared to U.S. national levels from the NHANES 2015-2016 sample (PFOS, PFOA, PFNA, PFHxS, PFHpS, PFDA, PFUnDA), and the 2017-2018 sample for Me-PFOSA, PFHPS using the Wilcoxon rank-sum test.

**Results:** Over 96% of SHOW participants had positive results for PFOS, PFHxS, PFHpS, PFDA, PFNA, and PFOA. In general, SHOW participants had lower serum levels across all PFAS when compared to NHANES. Serum levels increased with age and were higher among males and whites. These trends were seen in NHANES, except non-whites had higher PFAS levels at higher percentiles.

**Significance:** Wisconsin residents may have a lower overall body burden of some PFAS compounds compared to those seen by a nationally representative sample. Additional testing and characterization may be needed in Wisconsin, particularly among non-whites and low socioeconomic status, for which the SHOW sample had less representation compared to NHANES.

**Impact Statement:** The present study conducts biomonitoring of 38 PFAS in the state of Wisconsin and suggests that while most residents of Wisconsin have detectable levels of PFAS in their blood serum, they may have a lower body burden of some PFAS compared to a nationally representative sample. Older adults, males, and whites may have a higher body burden of PFAS relative to other groups both in Wisconsin and the wider United States.

## 1. Introduction

Per- and polyfluoroalkyl substances (PFAS) are a large family of manufactured, or human-made, fluorinated compounds produced for a variety of consumer products. PFAS are known as legacy chemicals because of their long half-life and ability to accumulate in the environment, as well as in animal and human blood serum and tissue [1-2]. The water and oil repellent properties of PFAS make them desirable for use as flame retardants and other common consumer products [2]. PFAS are highly stable compounds that not only repel water, oils, and lipids, but bind to proteins [3], and their strong carbon-fluorine bonds do not readily degrade. Therefore, some PFAS compounds can remain within all trophic levels for years or decades [4].

The stability of these compounds makes mitigation and removal of PFAS from the environment challenging. Environmental contamination sites have been identified for PFAS, which often arise from industrial production or agricultural practices using contaminated fertilizer [5]. Over time, PFAS leach into groundwater, surface waters, and soil. Major exposure pathways for PFAS in humans include consumption of contaminated food, especially fish and red meat [6]. Other sources of human exposure to PFAS include drinking water, contact with consumer products, and ingestion of dust particles [2].

Due to the widespread use of PFAS, recent environmental assessments have found them to be ubiquitous in the environment and in humans [7]. The U.S. Centers for Disease Control (CDC) biomonitoring of PFAS in the U.S. general population found nearly all the n=7991 U.S. residents tested via the 2011-2018 NHANES survey had detectable levels of one or more of the most studied long-chain PFAS: PFOA, PFOS, PFHxS, and PFNA [8]. This raised public health concerns, especially as PFAS research finds more associations between PFAS exposure and adverse human health effects [9]. Exposure to PFAS has been associated with cancer [10], higher cholesterol [11], liver enzyme changes [12], lower infant birth weight [13], decreased pediatric vaccine response [14], increased risk of gestational hypertension or pre-eclampsia [15], and immune suppression [16].

While PFOS and PFOA went largely unregulated for decades, in 2002 the U.S. Environmental Protection Agency (EPA) began to regulate PFAS, requiring manufacturers to notify the EPA of the manufacture or import of 13, and later 75, of the compounds [17]. Long-chain PFAS are still used in imported products from developing countries [18,19]. In June 2022, the EPA set a non-enforceable lifetime health advisory for PFOS and PFOA in drinking water to 0.02 and 0.004 parts per trillion, respectively [20]. A handful of states, including Wisconsin, have adopted the 2016 standard as the statewide regulatory standard for at least one PFAS compound [21]. This is a rapidly evolving regulatory landscape, with some states making recommendations more stringent than the EPA standard, and others making no recommendations at all [21]. However, there are an estimated 4000 PFAS compounds [17] produced by industry, no biomonitoring or epidemiological research on the vast majority of PFAS. Although long-chain PFASs (PFOA and PFAS) have been phased out of production in the U.S. and Canada, they are replaced by short-chain PFAS in industrial production. Additional biomonitoring and research are needed to understand how short-chain PFAS affect human health [22].

CDC biomonitoring of PFAS in the U.S. general population, and several other biomonitoring studies have emerged in the U.S. over the last decade. Little biomonitoring yet to be seen in the Upper Midwest and central region of the U.S. [23-27]. With NHANES data, the CDC established baseline exposure data for PFAS in the U.S., but the scale of the survey lacks the granularity needed to understand PFAS within smaller regions and demographic strata at the state or local level. The other community-based and localized biomonitoring studies to-date are not sampled in such a way to provide representation of PFAS exposure across an entire state. This study begins to fill this data gap by providing baseline PFAS levels representative of residents in the state of Wisconsin. The study uses serum samples from The Survey of the Health of Wisconsin (SHOW) cohort, the only statewide representative health survey in the U.S. modeled after NHANES.

In Wisconsin, PFAS have been found in groundwater supplies exceeding health-based recommendations [28]. Not only are there several contamination sites in Wisconsin, including the Marinette and Peshtigo areas in Marinette county due to production and testing of aqueous film-forming foams (AFFF) [29], but it’s history in paper production and known PCB and mercury contamination water ways from paper mills [30-32], in conjunction with agricultural use of wastewater sludge on crop fields, provide a landscape upon which residents may be at risk of PFAS exposure [33-34]. PFAS has been detected in not only groundwater, but in milk raised and produced in Wisconsin, the second largest dairy producing state [33]. Eighteen different PFAS compounds were detected in well samples in Madison, Wisconsin in domestic, municipal, and agricultural wells [35]. Furthermore, Wisconsin’s variable geography (forested, agricultural, and varying levels of urbanicity) and demographics are reasonably comparable to that of the broader United States in age structure, gender, and race [36].

The Wisconsin State Laboratory of Hygiene (WSLH) developed a new method to detect PFAS compounds at lower concentrations which increases the number of compounds available for assessment than have been used by many previous biomonitoring studies. In this study, we first examined an expanded list of PFAS compounds analyzed them using high-pressure liquid chromatography coupled with tandem mass spectrometric detection (HPLC-MS/MS) and, where we were able to, compared SHOW PFAS serum levels to those PFAS serum levels of a nationally representative sample of the United States (NHANES). While other studies have drawn comparisons to NHANES PFAS levels, to our knowledge, no other statewide representative samples have compared PFAS levels to NHANES.

## 2. Methods

### 2.1 Survey of the Health of Wisconsin (SHOW)

Survey data and biospecimens came from the 2014-2016 Survey of the Health of Wisconsin (SHOW) statewide representative sample of adults ages 18 and older (n=1957). Details on the SHOW sampling frame, recruitment, and methods are described elsewhere [37]. In brief, the SHOW 2014–2016 cohort was designed as a three-year sample. A three-stage cluster-sampling approach was employed. One county per strata was randomly selected within the strata of county mortality rates, followed by random selection of census block groups by poverty status strata. Then 30–35 residential households were randomly selected via US postal service listings. Housed within the Department of Population Health Sciences at the University of Wisconsin-Madison, SHOW conducts statewide population–based recruitment and longitudinal follow-up of a representative sample of state residents SHOW is unique in that it is the only statewide household-based examination survey in the United States and offers an abundance of survey and physical measurement data upon which to characterize PFAS exposure within our sample.

Modeled after the CDC’s NHANES, its survey data includes demographics (age, gender, education, income, race), behaviors (smoking, diet), occupation (military service, firefighter), housing (type and age of residence), drinking water characteristics (private well vs. municipal, depth of private well, consumption pattern, treatment/filter use), diet (fish, dairy) and chronic health conditions (cancer, diabetes, hypertension, obesity). In addition, participant households span rural, urban and suburban settings and are geocoded to allow for linkages to contextual environmental data, including potential PFAS sources (landfills, industrial and municipal wastewater facilities, agricultural fields).

Households are randomly selected from commercially available US Postal Service residential address listings using a random probability-based sampling procedure and SHOW trained interviewers conduct in-home visits upon which they gather health information via computer assisted personal interview (CAPI). Participants also complete a self-administered paper questionnaire. Following an in-home visit, participants visit a collection site near their home where phlebotomists measure blood pressure, weight, height, waist and hip circumference, respiratory function, and collect venous blood and urine samples. Several tubes of venous blood (about 55–60 ml in total) are immediately processed for serum and plasma, aliquoted into cryovials and frozen at −80C. SHOW participants consent to the use of biospecimens for unspecified research. The core SHOW study and this study is approved by the University of Wisconsin Health Sciences Institutional Review Board and all biosecurity and institutional safety procedures are HIPAA compliant.

For this study, a subset of (n=605) participants were selected at random from SHOW’s 2014– 2016 statewide representative sample. For this retrospective sample analysis, a single random sample of adults 18+ (n=50; stratified by race, sex, age), was pulled from each of 12 counties representing each of 5 health regions in the state. Only individuals with stored serum were included when selecting the study sample. This allows for the assessment of historic PFAS exposures across Wisconsin and accelerates the ability to observe time-dependent trends in exposures when prospective sampling is performed. An additional sample was pulled per health region as substitution in case any samples were not viable for PFAS analyses. Serum samples were extracted from SHOW’s freezer and sent to the Wisconsin State Laboratory of Hygiene on dry ice for PFAS analyses.

### 2.3 PFAS Sample Analysis

The PFAS analyses were performed by the Wisconsin State Lab of Hygiene (WSLH) using high-pressure liquid chromatography coupled with tandem mass spectrometric detection (HPLC-MS/MS). The 38 PFAS compounds tested were selected based on several considerations, including, (1) cover the spectrum of compounds to look for class-specific bioaccumulation, (2) include compounds most often used in products and tested by CDC in NHANES, (3) include both short and long chain PFAS to assess differences by type, (4) include emerging compound classes for which we have little data, (5) consider potential toxicity estimates, and (6) limited by the availability of suitable materials to assure reliable measurement.

The WSLH test method was adapted primarily from a method developed by Minnesota Dept. of Public Health [38], with elements from CDC Method 6304.08 [39], the New York State Dept. of Health [40], and the Michigan Dept. of Community Health [41]. Several of these documents are laboratory manuals, some shared privately between colleagues. Sample preparation involved spiking aliquots of serum with an isotopically labeled PFAS mixture. Acetonitrile was added to precipitate proteins, followed by vortex mixing. Samples were then centrifuged, the supernatant transferred to a 96-well plate, and evaporated to a volume of 100-200 mL.

Prepared samples were injected on an Agilent 1290 Infinity UPLC (Santa Clara, CA) equipped with a BEH C18 1.7mm 2.1×50mm Column (Waters, Milford, MA). Good chromatographic separation was achieved using a reverse phase gradient, with a 20 minute run time at 45°C. Sample detection and quantification was achieved with a Sciex 4500 MS/MS system (Toronto, CA), employing multiple reaction mode (MRM) scanning with negative polarity turbospray ionization. Q1 and Q3 masses, declustering Potential (DP), collision energy (CE) and collision cell exit potential (CXP) values for each MRM transition were optimized by parameter ramping experiments with direct standard solution infusion to the source. Sciex Analyst version 1.6.3 was used for data acquisition and results calculation.

Method quality control characteristics included a method blank and seven standard linear calibration standards (r ≥0.995). Standards were verified using a second source material. Analytical signal/noise for S_1_ was required to be ≥10. Ion confirmation ratio for samples was required to fall within ±30% of the mean ratio of the standards, with exceptions for PFOS, FOSA, PFDA, PFNS, 11Cl-PF3OUdS, PFHxA, PFTrDA, PFDS, PFTeDA, and 8:2 diPAP, which were widened to ±40% based on poor confirmation ion sensitivity. Method detection limits varied considerably by compound, as did the upper quantification limits, but compare favorably with NHANES quantification thresholds for those compounds. Three levels of analytical controls were measured in every analytical run, and acceptable control values bracketed all test samples. Method validation also included precision, analytical measurement range, and spike recovery assessment. Additional details on the analytical method area and observations on compounds not comparable with NHANES are in preparation.

### 2.4 National Health and Nutrition Examination Survey (NHANES)

NHANES is an ongoing national cross-sectional survey administered by the National Center of Health Statistics (NCHS) within the CDC. NHANES uses a multistage cluster sampling approach to examine a study population that is representative of the United States’ non-institutionalized population. Blood samples are taken from all NHANES participants 12 years of age and older. Approximately 2,000 of these samples were analyzed for several PFAS compounds in each cycle. The NHANES 2015-2016 sample intentionally oversamples Hispanic, Non-Hispanic black, Non-Hispanic Asian, Non-Hispanic white and others at or below 185 percent of the Department of Health and Human Services (HHS) poverty guidelines, and Non-Hispanic white and other people aged 80 years and older. NHANES releases publicly available laboratory and demographic data on these participants. Education and smoking status for adolescents (age 12-19) is considered sensitive data and not included in public use datasets. Documentation for NHANES data includes detailed laboratory methods [42]. For this study, SHOW study sample PFAS serum levels were compared with those from the National Health and Nutrition Examination Survey (NHANES) 2015-2016 sample (n=1829) for all corresponding samples. However, given that NHANES did not sample the entire suite of compounds in this study, the 2017-2018 (n=1862) samples were compared for select compounds (ME-PFOSA and PFHPS).

### 2.5 Demographics

Self-reported demographics data for gender, age, race/ethnicity, highest education level attained, and income were collected by CAPI. Smoking status data were obtained through a self-administered questionnaire (SAQ) for the Wisconsin Sample and personal interview for NHANES. Income/poverty ratio was calculated by dividing the midpoint of reported household income range by HHS poverty guidelines, which is calculated by the number of people supported by that income [43]. Body mass index (BMI) (kg/m^2^) was calculated using WHO standards by dividing measured height (cm) by 100, and squaring that value, and then dividing weight (kg) by that value. SHOW and NHANES participants were stratified by gender, age, and race for analysis of PFAS serum levels. Age groups were determined by generational changes (18-39, 40-59, 60+), and race was grouped by non-Hispanic white and non-white. Minors in the NHANES samples were excluded from the final analysis.

### 2.6 Statistical Analyses

All statistical analyses were performed using SAS v9.4. All thirty-eight PFAS compounds were reported for SHOW using the LLOD available from the WSLH assays. However, only compounds analyzed in both SHOW and available in NHANES public use laboratory data were included in comparative data analysis, and only compounds with more than 15% positivity in the SHOW sample could be reliably compared to NHANES. Thus, weighted geometric means for 8 PFAS compounds, as well as weighted geometric means for the 50^th^, 75^th^, 90^th^, and 95^th^ percentiles and their corresponding 95% confidence intervals were calculated for SHOW and NHANES (see Table 2 for complete list of PFAS). The lower limit of detection (LLOD) for all compounds in NHANES was 0.1 ng/mL, with values lower than the LLOD set to 0.1 divided by the square root of 2, or approximately 0.07. The LLODs for SHOW participants were different from 0.1, but to make direct comparisons, all PFAS compounds within SHOW were assigned the same LLOD as NHANES for comparison analyses. Therefore, weighted geometric means differ for SHOW in the comparison tables with NHANES, when compared to findings from SHOW with the LLOD from WSLH, unaltered to match NHANES.

**Table 1.**
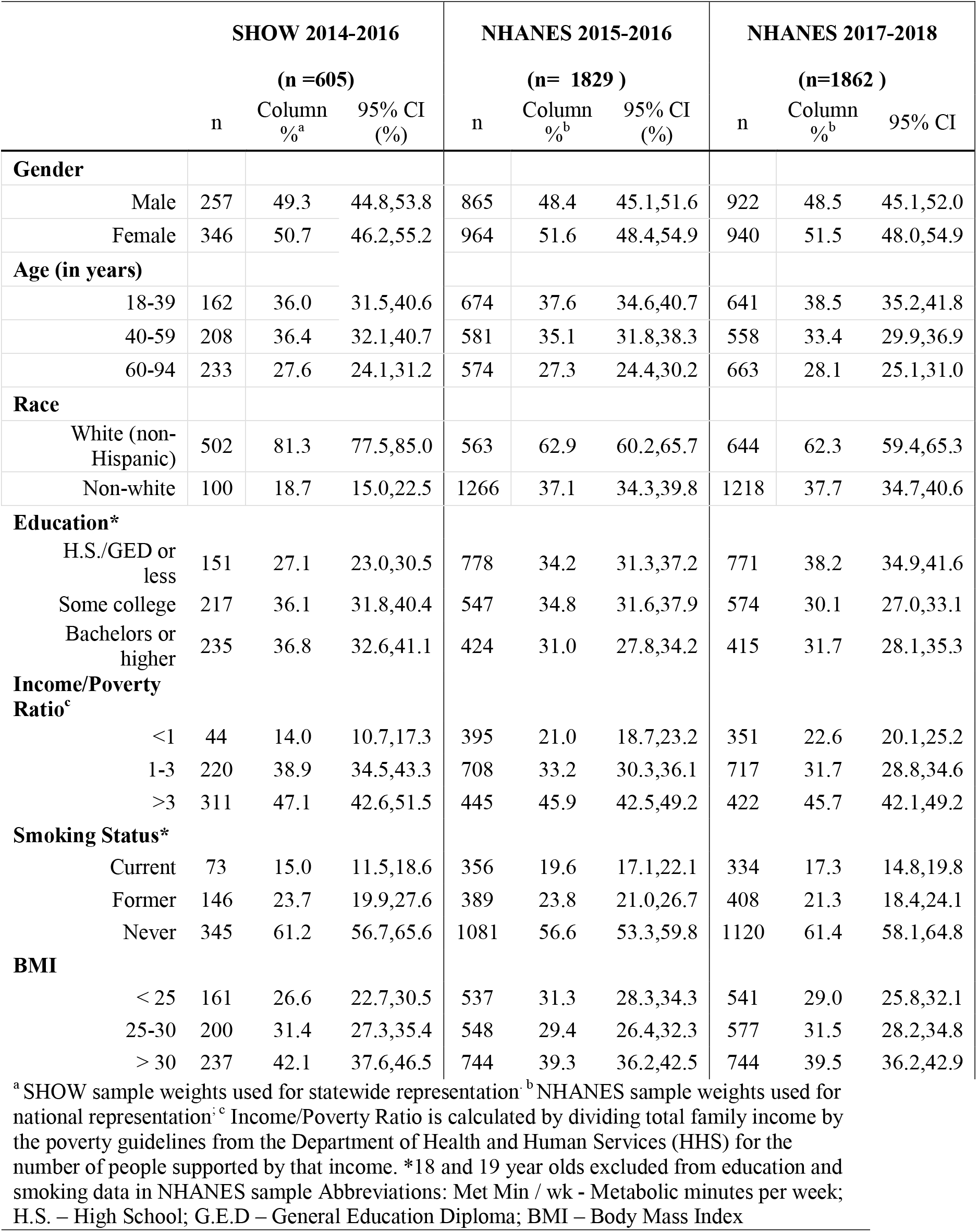
Demographics characteristics of SHOW 2014-2016 compared with NHANES 2015-2016, and NHANES 2017-2018 cohorts.

| SHOW 2014-2016 |  |  |  | NHANES 2015-2016 |  |  | NHANES 2017-2018 |  |  |
| --- | --- | --- | --- | --- | --- | --- | --- | --- | --- |
| (n =605) |  |  |  | (n= 1829 ) |  |  | (n=1862 ) |  |  |
|  | n | Column %<sup>a</sup> | 95% CI (%) | n | Column %<sup>b</sup> | 95% CI (%) | n | Column %<sup>b</sup> | 95% CI |
| **Gender** |  |  |  |  |  |  |  |  |  |
| Male | 257 | 49.3 | 44.8,53.8 | 865 | 48.4 | 45.1,51.6 | 922 | 48.5 | 45.1,52.0 |
| Female | 346 | 50.7 | 46.2,55.2 | 964 | 51.6 | 48.4,54.9 | 940 | 51.5 | 48.0,54.9 |
| **Age (in years)** |  |  |  |  |  |  |  |  |  |
| 18-39 | 162 | 36.0 | 31.5,40.6 | 674 | 37.6 | 34.6,40.7 | 641 | 38.5 | 35.2,41.8 |
| 40-59 | 208 | 36.4 | 32.1,40.7 | 581 | 35.1 | 31.8,38.3 | 558 | 33.4 | 29.9,36.9 |
| 60-94 | 233 | 27.6 | 24.1,31.2 | 574 | 27.3 | 24.4,30.2 | 663 | 28.1 | 25.1,31.0 |
| **Race** |  |  |  |  |  |  |  |  |  |
| White (non-Hispanic) | 502 | 81.3 | 77.5,85.0 | 563 | 62.9 | 60.2,65.7 | 644 | 62.3 | 59.4,65.3 |
| Non-white | 100 | 18.7 | 15.0,22.5 | 1266 | 37.1 | 34.3,39.8 | 1218 | 37.7 | 34.7,40.6 |
| **Education\*** |  |  |  |  |  |  |  |  |  |
| H.S./GED or less | 151 | 27.1 | 23.0,30.5 | 778 | 34.2 | 31.3,37.2 | 771 | 38.2 | 34.9,41.6 |
| Some college | 217 | 36.1 | 31.8,40.4 | 547 | 34.8 | 31.6,37.9 | 574 | 30.1 | 27.0,33.1 |
| Bachelors or higher | 235 | 36.8 | 32.6,41.1 | 424 | 31.0 | 27.8,34.2 | 415 | 31.7 | 28.1,35.3 |
| **Income/Poverty Ratio<sup>c</sup>** |  |  |  |  |  |  |  |  |  |
| <1 | 44 | 14.0 | 10.7,17.3 | 395 | 21.0 | 18.7,23.2 | 351 | 22.6 | 20.1,25.2 |
| 1-3 | 220 | 38.9 | 34.5,43.3 | 708 | 33.2 | 30.3,36.1 | 717 | 31.7 | 28.8,34.6 |
| >3 | 311 | 47.1 | 42.6,51.5 | 445 | 45.9 | 42.5,49.2 | 422 | 45.7 | 42.1,49.2 |
| **Smoking Status\*** |  |  |  |  |  |  |  |  |  |
| Current | 73 | 15.0 | 11.5,18.6 | 356 | 19.6 | 17.1,22.1 | 334 | 17.3 | 14.8,19.8 |
| Former | 146 | 23.7 | 19.9,27.6 | 389 | 23.8 | 21.0,26.7 | 408 | 21.3 | 18.4,24.1 |
| Never | 345 | 61.2 | 56.7,65.6 | 1081 | 56.6 | 53.3,59.8 | 1120 | 61.4 | 58.1,64.8 |
| **BMI** |  |  |  |  |  |  |  |  |  |
| < 25 | 161 | 26.6 | 22.7,30.5 | 537 | 31.3 | 28.3,34.3 | 541 | 29.0 | 25.8,32.1 |
| 25-30 | 200 | 31.4 | 27.3,35.4 | 548 | 29.4 | 26.4,32.3 | 577 | 31.5 | 28.2,34.8 |
| > 30 | 237 | 42.1 | 37.6,46.5 | 744 | 39.3 | 36.2,42.5 | 744 | 39.5 | 36.2,42.9 |

<sup>a</sup> SHOW sample weights used for statewide representation; <sup>b</sup> NHANES sample weights used for national representation; <sup>c</sup> Income/Poverty Ratio is calculated by dividing total family income by the poverty guidelines from the Department of Health and Human Services (HHS) for the number of people supported by that income. \*18 and 19 year olds excluded from education and smoking data in NHANES sample Abbreviations: Met Min / wk - Metabolic minutes per week; H.S. – High School; G.E.D – General Education Diploma; BMI – Body Mass Index

**Table 2.**
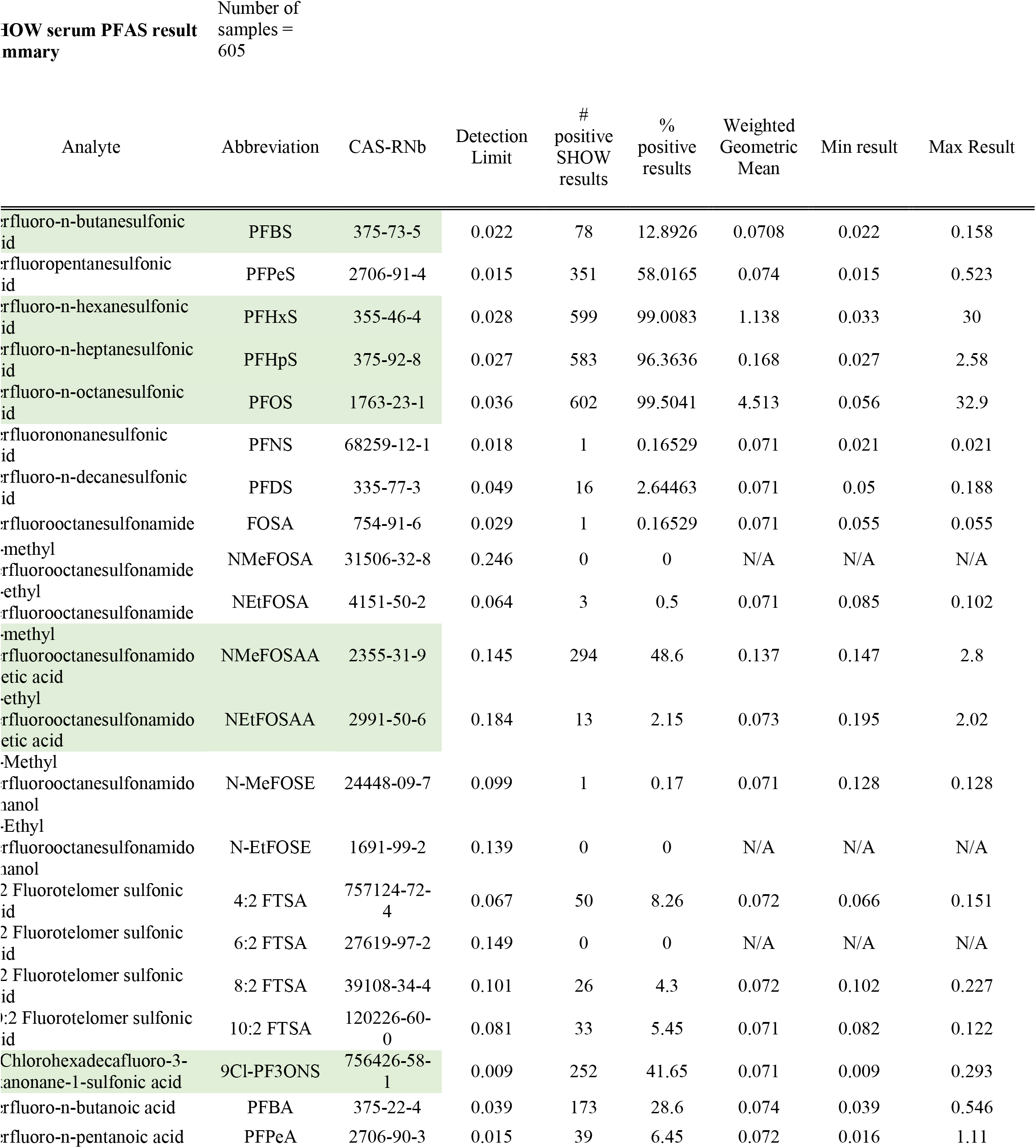
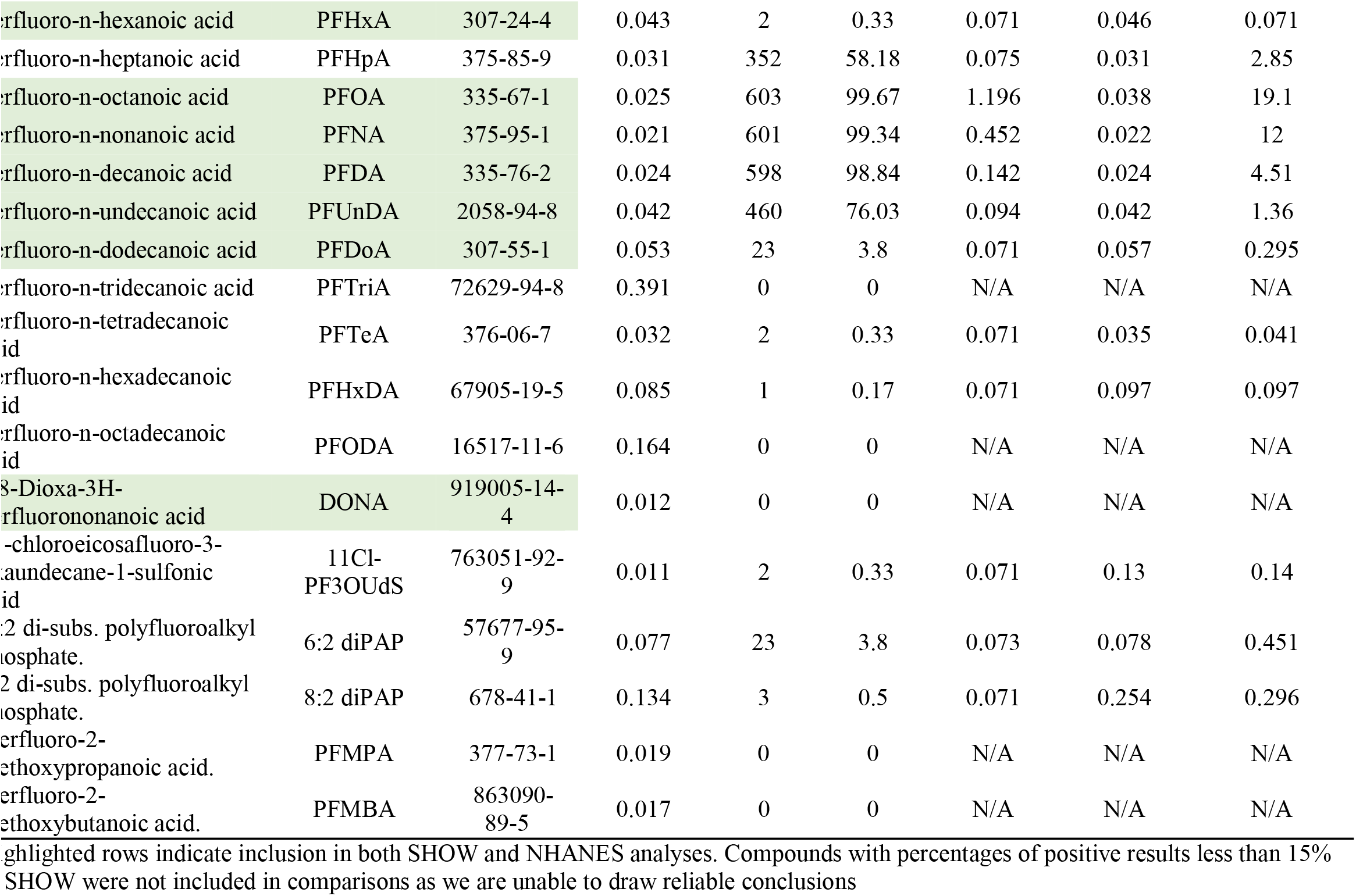
Summary of SHOW results for all PFAS analytes

PFOS and PFOA were analyzed as the sum of their linear and branched isomers, consistent with NHANES methods. Data from minors ages 12-17 in the NHANES sample were used to validate data analysis methods against published NHANES data tables, but minors were not included in the final analysis as all SHOW participants were over the age of 18. Geometric means of serum levels and corresponding 95% confidence intervals were calculated for all compounds and compared between SHOW and NHANES. Both NHANES and SHOW geometric means were calculated using subsample weights, and domains of gender, age, and race.

While overall and demographic strata comparisons of PFAS serum levels in SHOW vs. NHANES are weighted, due to the different complexities of sampling in both SHOW and NHANES, unweighted comparisons of PFAS serum concentrations were evaluated using the Wilcoxon rank-sum test to test difference. Compounds detected above the limit of detection in fewer than 50% of individuals were not included in further analysis comparisons with NHANES.

## 3. Results

### 3.1 Study Sample

Descriptive characteristics of the SHOW 2014-2016 subsample compared with NHANES 2015-2016 and NHANES 2017-2018 samples are presented in Table 1. The SHOW study sample is comparable to NHANES in terms of gender and age distribution. There are slightly more females (50.7%) than males in SHOW; similar in NHANES (51.5-51.6%). The SHOW sample differs from NHANES most in terms of racial diversity. The SHOW consists of 81.3% non-Hispanic white, much higher when compared with NHANES where 62.3-62.9% are non-Hispanic white. SHOW also had slightly more participants with >3 income to poverty ratio, fewer smokers, and more with a BMI > 30 (Table 1). SHOW participants also tended to be more educated than NHANES participants on average, with 27.1% of SHOW participants reporting high school/GED or less in SHOW, compared with 34.2% in NHANES.

### 3.2 Prevalence of detectable serum PFAS levels

Table 2 depicts the detection limit and summary statistics for the number of positive results, geometric mean, minimum and maximums for all 38 PFAS compounds. Nine of the 38 PFAS compounds were detected in at least 50% of SHOW participants (PFOS, PFOA, PFNA, PFHxS, PFDA, PFHpS, PFUnDA, PFHpA, PFPeS) (Table 2). More than 96% of SHOW participants had serum levels above the lower limit of detection for six PFAS analytes, PFOS, PFOA, PFNA, PFHxS, PFDA, and PFHpS with geometric mean values being 4.513, 1.196, 0.452, 1.138, 0.142, and 0.168 ng/mL, respectively. Eight PFAS compounds were below the limit of detection in 100% of the study sample (NMeFOSA, N-EtFOSE, 6:2 FTSA, PFTriA, PFODA, DONA, PFMPA, PFMBA). SHOW participants had much higher levels of PFOS compared to other compounds, with a whole sample geometric mean of 4.513 μg/L (Table 2).

### 3.3 PFAS Comparison of SHOW to NHANES by demographics

Table 3 depicts comparisons in geometric means and percentiles between the entire SHOW and NHANES samples where LLOD has been adjusted for SHOW to match NHANES. Tables 4-6 (and supplementary tables 1-5) depict geometric means and percentiles comparing SHOW and NHANES by demographic strata. Weighted geometrics means of PFAS serum levels were slightly higher among NHANES study sample compared to SHOW for PFOS, PFOA, PFNA, PFHxS, PFDA, and PFHPS. However, only PFOA and PFNA were statistically higher among NHANES compared SHOW (p <0.001). Most notable differences in weighted geometric means were seen for PFHPS, PFOA, and PFNA, where NHANES serum samples were 35.3%, 32.5%, and 31.1% higher than in SHOW, respectively. PFDA, PFOS, and PFHxS weighted geometrics mean serum levels were 14.3%, 10.2%, 7.0% higher in NHANES when compared to SHOW. PFOS weighted geometric means (95% CI) were highest of all PFAS compounds in both study samples; 4.51 (4.18-4.87) in SHOW compared to 4.97 (4.71-5.25) in NHANES.

**Table 3.**
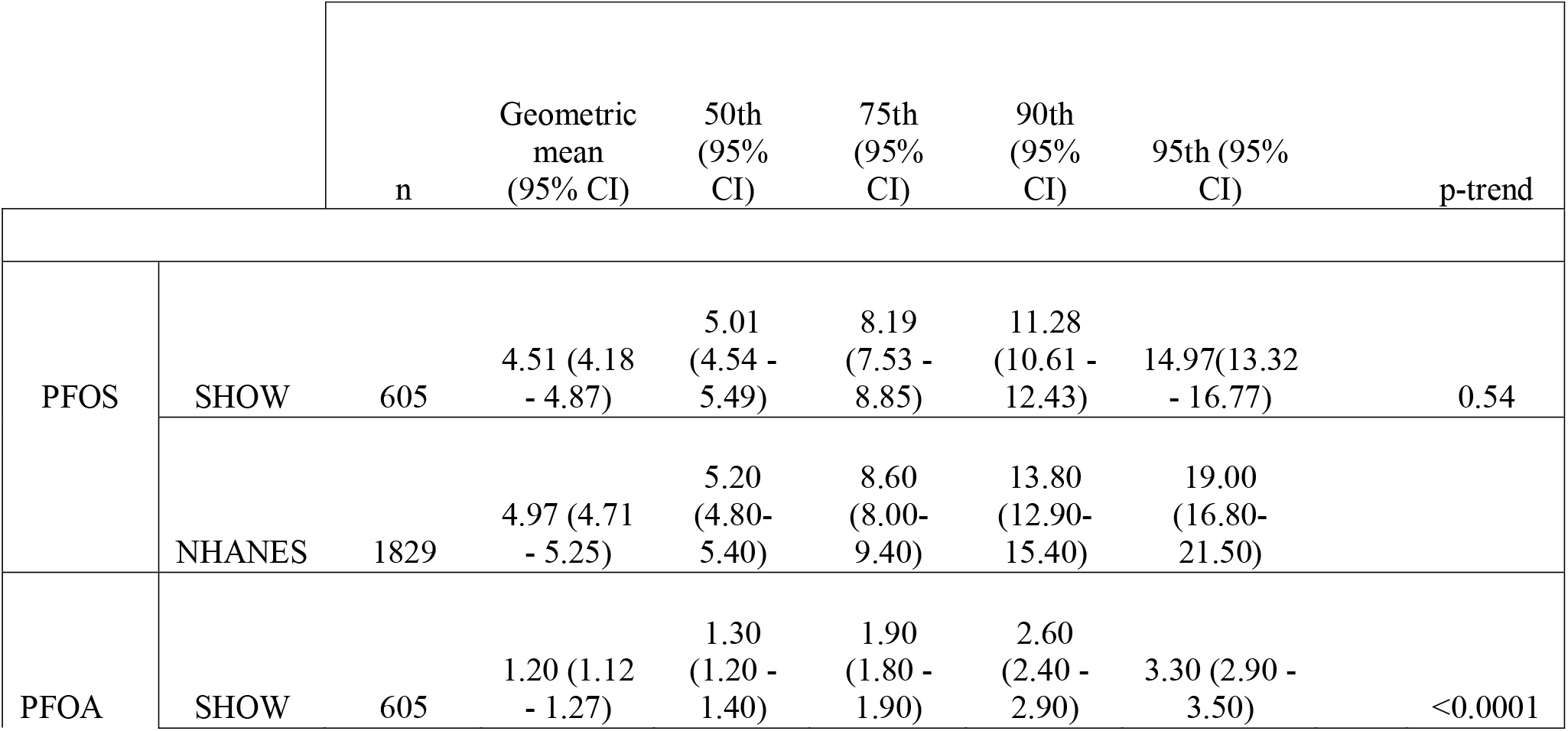
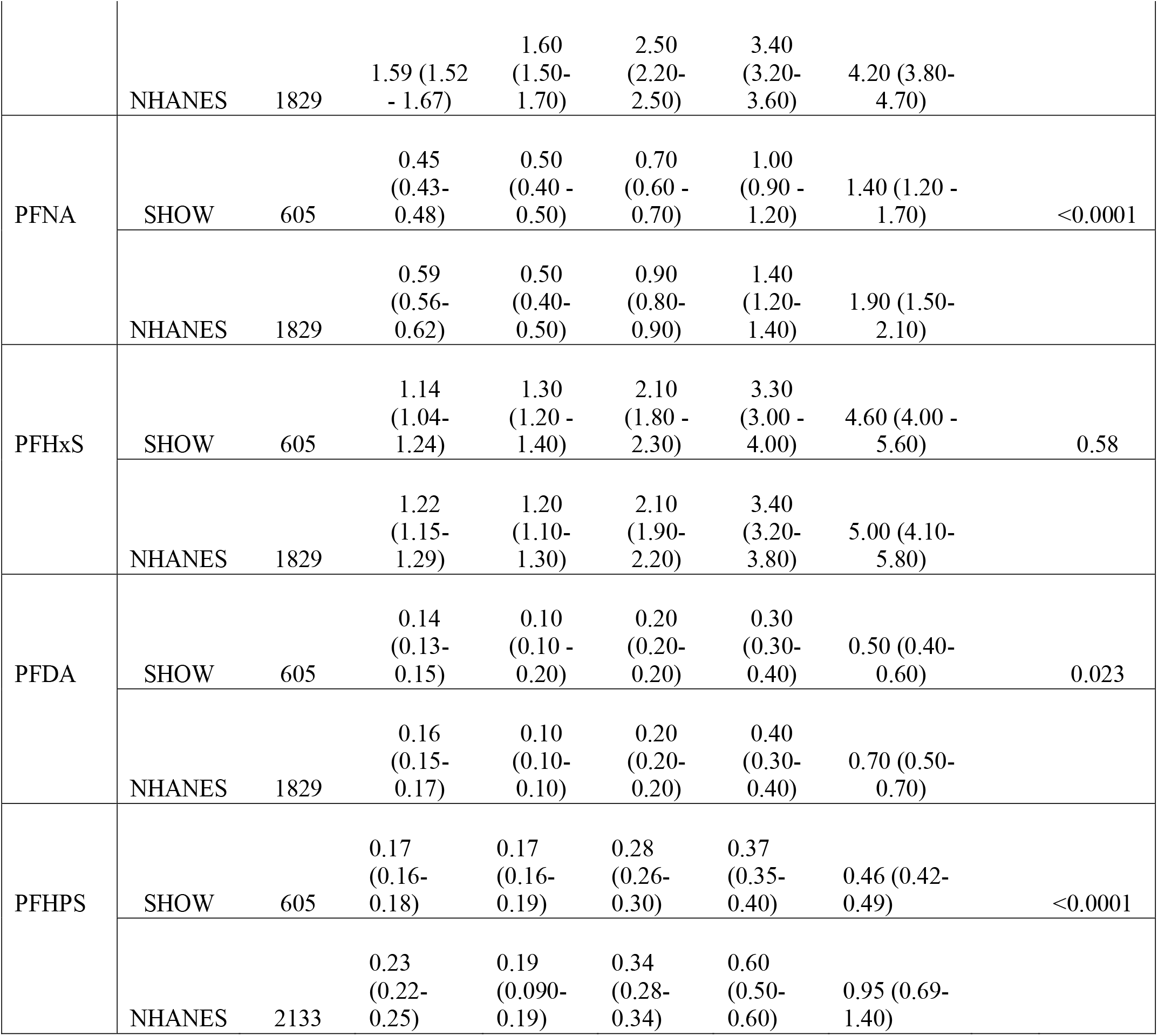
Geometric Means and Percentile Comparisons between SHOW and NHANES

**Table 4.**
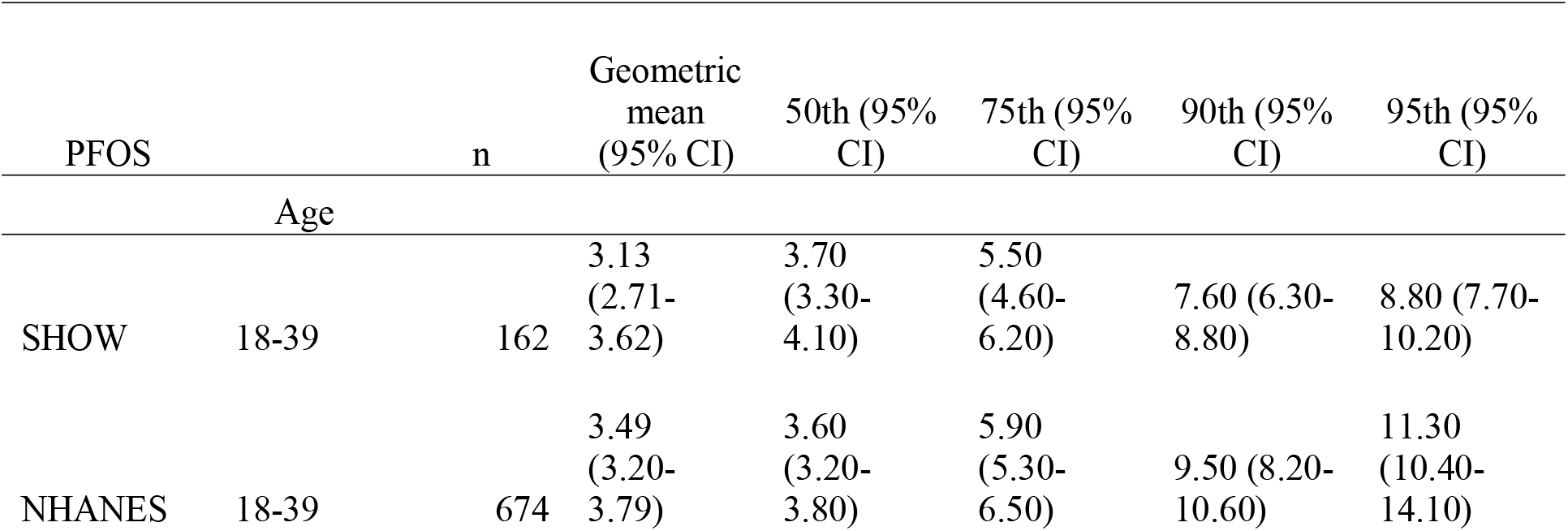
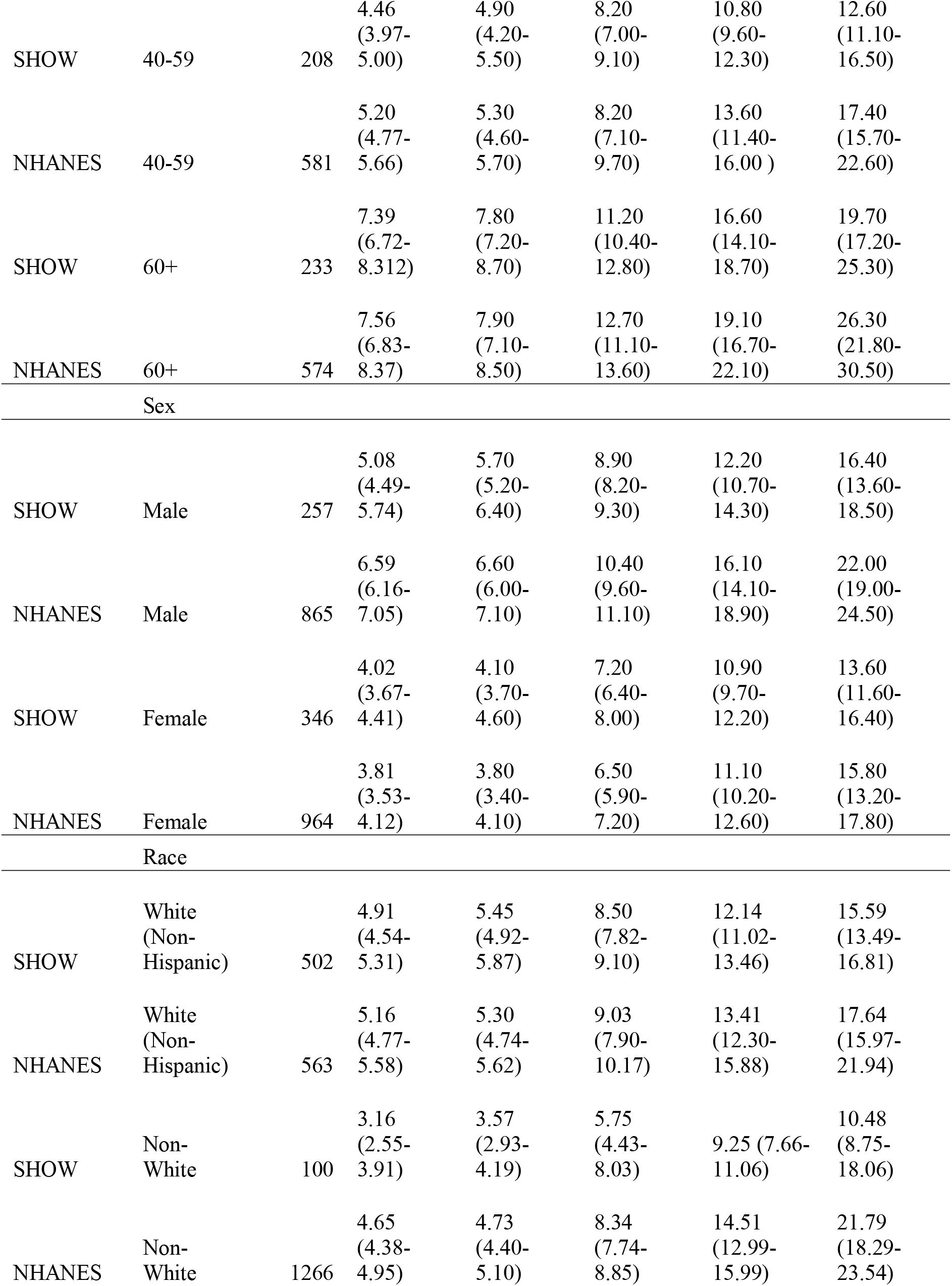
Geometric means and percentiles of serum PFOS by demographic characteristcs-comparing 18+ in SHOW 2014-2016 with U.S. NHANES 2015-2016

**Table 5.** Geometric means and percentiles of serum PFOA by demographic characteristics-comparing 18+ in SHOW 2014-2016 with U.S. NHANES 2015-2016

| PFOA |  | n | Geometric<br>mean (95%<br>CI) | 50th (95%<br>CI) | 75th (95%<br>CI) | 90th (95%<br>CI) | 95th (95%<br>CI) |
| --- | --- | --- | --- | --- | --- | --- | --- |
| Age |  |  |  |  |  |  |  |
| SHOW | 18-39 | 162 | 0.93 (0.82-1.05) | 1.10 (0.90-1.20) | 1.60 (1.40-1.80) | 2.00 (1.90-2.30) | 2.40 (2.00-3.20) |
| NHANES | 18-39 | 674 | 1.31 (1.21-1.40) | 1.40 (1.20-1.40) | 2.00 (1.80-2.20) | 2.80 (2.50-3.00) | 3.50 (2.90-3.90) |
| SHOW | 40-59 | 208 | 1.28 (1.16-1.41) | 1.30 (1.20-1.50) | 1.80 (1.70-2.10) | 2.60 (2.40-3.10) | 3.30 (2.70-4.60) |
| NHANES | 40-59 | 581 | 1.64 (1.53-1.77) | 1.60 (1.40-1.80) | 2.40 (2.10-2.50) | 3.30 (3.10-3.60) | 3.90 (3.50-5.20) |
| SHOW | 60+ | 233 | 1.51 (1.38-1.65) | 1.50 (1.40-1.80) | 2.30 (2.10-2.50) | 3.30 (2.80-3.50) | 3.50 (3.40-4.20) |
| NHANES | 60+ | 574 | 2.01 (1.81-2.22) | 2.10 (1.90-2.20) | 2.80 (2.60-3.10) | 4.20 (3.60-4.90) | 5.20 (4.60-6.90) |
| Sex |  |  |  |  |  |  |  |
| SHOW | Male | 257 | 1.25 (1.13-1.37) | 1.40 (1.30-1.50) | 1.90 (1.80-2.00) | 2.60 (2.40-3.00) | 3.20 (2.90-3.60) |
| NHANES | Male | 865 | 1.85 (1.75-1.96) | 1.90 (1.70-2.00) | 2.60 (2.50-2.70) | 3.40 (3.20-3.60) | 4.10 (3.60-5.010) |
| SHOW | Female | 346 | 1.15 (1.06-1.25) | 1.20 (1.10-1.20) | 1.80 (1.70-2.00) | 2.70 (2.40-3.10) | 3.50 (2.90-4.20) |
| NHANES | Female | 964 | 1.39 (1.29-1.49) | 1.40 (1.30-1.50) | 2.30 (2.00-2.40) | 3.40 (2.90-3.90) | 4.30 (3.90-6.10) |
| Race |  |  |  |  |  |  |  |
| SHOW | White<br>(Non-Hispanic) | 502 | 1.28 (1.20-1.37) | 1.40 (1.30-1.50) | 1.90 (1.80-2.00) | 2.80 (2.50-3.10) | 3.40 (3.10-4.00) |
| NHANES | White<br>(Non-Hispanic) | 563 | 1.72 (1.60-1.84) | 1.80 (1.60-1.90) | 2.60 (2.40-2.70) | 3.50 (3.20-3.80) | 4.60 (3.90-5.60) |
| SHOW | Non-White | 100 | 0.90 (0.77-1.05) | 1.00 (0.80-1.10) | 1.40 (1.20-1.80) | 2.00 (1.60-2.50) | 2.40 (2.00-3.00) |
| NHANES | Non-White | 1266 | 1.40 (1.34-1.47) | 1.40 (1.30-1.40) | 2.10 (2.00-2.20) | 3.10 (2.80-3.30) | 4.00 (3.60-4.10) |

**Table 6.** Geometric means and percentiles of serum PFNA by demographic characteristcs-comparing 18+ in SHOW 2014-2016 with U.S. NHANES 2015-2016

| PFNA |  | n | Geometric mean (95% CI) | 50th (95% CI) | 75th (95% CI) | 90th (95% CI) | 95th (95% CI) |
| --- | --- | --- | --- | --- | --- | --- | --- |
| Age |  |  |  |  |  |  |  |
| SHOW | 18-39 | 162 | 0.36 (0.32-0.41) | 0.40 (0.30-0.40) | 0.60 (0.50-0.70) | 0.80 (0.70-1.10) | 1.10 (0.80-1.40) |
| NHANES | 18-39 | 674 | 0.46 (0.43-0.50) | 0.40 (0.30-0.40) | 0.70 (0.60-0.70) | 1.00 (0.90-1.00) | 1.30 (1.10-1.50) |
| SHOW | 40-59 | 208 | 0.44 (0.40-0.48) | 0.40 (0.40-0.50) | 0.60 (0.60-0.70) | 0.90 (0.80-1.20) | 1.30 (1.00-1.70) |
| NHANES | 40-59 | 581 | 0.60 (0.55-0.66) | 0.50 (0.50-0.60) | 0.90 (0.80-1.00) | 1.30 (1.20-1.50) | 1.90 (1.40-2.30) |
| SHOW | 60+ | 233 | 0.62 (0.56-0.67) | 0.60 (0.60-0.70) | 0.90 (0.80-0.90) | 1.40 (1.20-1.80) | 2.10 (1.50-2.60) |
| NHANES | 60+ | 574 | 0.78 (0.71-0.85) | 0.70 (0.60-0.80) | 1.10 (1.00-1.30) | 1.70 (1.50-2.00) | 2.30 (1.90-2.60) |
| Sex |  |  |  |  |  |  |  |
| SHOW | Male | 257 | 0.47 (0.43-0.51) | 0.50 (0.40-0.50) | 0.70 (0.60-0.80) | 1.00 (0.90-1.20) | 1.40 (1.10-2.10) |
| NHANES | Male | 865 | 0.64 (0.60-0.69) | 0.50 (0.40-0.50) | 0.90 (0.80-1.00) | 1.40 (1.10-1.40) | 1.90 (1.50-2.10) |
| SHOW | Female | 346 | 0.44 (0.40-0.48) | 0.40 (0.40-0.50) | 0.70 (0.60-0.80) | 1.10 (0.90-1.30) | 1.40 (1.20-2.00) |
| NHANES | Female | 964 | 0.54 (0.50-0.58) | 0.40 (0.30-0.40) | 0.80 (0.70-0.80) | 1.30 (1.10-1.40) | 1.90 (1.40-2.40) |
| Race |  |  |  |  |  |  |  |
| SHOW | White (Non-Hispanic) | 502 | 0.48 (0.45-0.51) | 0.50 (0.40-0.50) | 0.70 (0.70-0.80) | 1.10 (0.90-1.30) | 1.50 (1.30-2.10) |
| NHANES | White (Non-Hispanic) | 563 | 0.58 (0.54-0.62) | 0.50 (0.40-0.50) | 0.90 (0.70-0.90) | 1.30 (1.10-1.40) | 1.90 (1.40-2.30) |
| SHOW | Non-White | 100 | 0.36 (0.31-0.41) | 0.30 (0.30-0.40) | 0.60 (0.50-0.70) | 0.80 (0.60-1.10) | 1.00 (0.80-1.50) |
| NHANES | Non-White | 1266 | 0.60 (0.57-0.63) | 0.50 (0.40-0.50) | 0.90 (0.80-1.00) | 1.40 (1.30-1.50) | 1.90 (1.60-2.00) |

Serum levels increased with age in both SHOW and NHANES for all 6 PFAS compounds. In SHOW, 18–39-year-olds had a PFOS geometric mean of 3.13 (2.71-3.62) compared to 60+ year-olds 7.39 (6.72-8.31); whereas the 95^th^ percentile was 8.80 (7.70-10.20) for 18–39-year-olds and 26.3 (21.8-30.5) for 60+ year-olds. Similar trends held for NHANES.

In SHOW, males had higher serum levels of PFOS, PFOA, PFNA, PFHxS, and PFHPS than females; a difference in weighted geometric mean of 1.06, 0.1, 0.03, and 0.5 ng/mL, respectively. Levels did not differ by sex for PFDA; these trends by sex were seen in both the NHANES and SHOW samples for all PFAS.

Weighted geometric mean serum levels were higher among whites (non-Hispanic) in SHOW and NHANES for PFOA, PFOS, PFHxS and PFHPS when compared to nonwhites. These trends held in the 50-95th percentiles for SHOW, but for NHANES, these trends were opposite at the higher percentiles (90^th^ and 95^th^) for PFHPS and PFOS. PFHPS was 40% higher, and PFOS was 20% higher, among nonwhites compared to whites (non-Hispanic) in NHANES at the 95th percentile.

SHOW and NHANES samples were largely comparable for other PFAS compounds, (see Supp. Tables 1-5), albeit low levels of detection were seen for both SHOW and NHANES for these additional PFAS compounds.

## 4. Discussion

To our knowledge this is the first study to characterize PFAS serum levels among a statewide representative sample and compare levels to those of NHANES. This study expanded biomonitoring to a panel of 38 PFAS compounds and detected concentrations in serum with LLOD lower than many prior studies. The study leveraged the SHOW cohort, the only statewide adult cohort in the U.S. modeled after NHANES to provide baseline data for the state of Wisconsin. Beyond NHANES, current understanding of human serum PFAS levels come from specific sub-population cohorts (i.e. occupational, maternal-child cohorts) [23,44], high-risk or localized communities near contamination [24-26]. While baseline PFAS levels are known at a national scale using NHANES, those data lack the granularity needed at the state level and within subregions and strata in a state. SHOW’s statewide cohort provides PFAS serum prevalence for the state, and within subpopulations and communities, spanning different demographics, socio-economic backgrounds, and neighborhood environments. This study fills a data gap and allows for identification of previously unknown high-risk areas or sub-populations upon which researchers and state health officials can target additional biomonitoring and testing.

Among the 38 PFAS analytes tested, six were widely prevalent among SHOW; more than 96% of SHOW participants had detectable serum levels of PFOS, PFOA, PFNA, PFHxS, PFDA, and PFHpS. These findings align with what other studies have found. For example, Yu and colleagues conducted statewide testing in New Jersey from remnant sera from clinical labs and blood banks (n=1030) and of the 12 PFAS compounds tested, PFOA, PFNA, PFOS, and PFHxS were the ones detected in over 99% of the study population with geometrics means greater than 0.5 μg/L [44]. Even among high-risk or communities near contamination sites, the same PFAS compounds were the most widespread, and with the highest geometric means. Among n=192 claimants from class-action lawsuit in Paulsboro, New Jersey, who lived near a contamination site, of 13 PFAS compounds tested, PFOS, PFOA, PFNA and PFHxS had the highest prevalence, with over 70% with detectable levels [24]. This held true in other states, such as the communities tested near military bases in Pennsylvania [45] where PFOS, PFOA, PFHxS, and PFNA were highest and most prevalent of the 11 PFAS tested, and those in the Annison Community Health Survey living near a high-risk manufacturing site in Alabama, where among the 8 PFAS tested, PFOS, PFNA, PFOA, and PFHxS were detected in >96% [26].

Geometric means and trends cross demographic strata and percentiles among SHOW were like those seen in NHANES. Yet, NHANES consistently had slightly higher geometric means for all 6 main PFAS compounds analyzed (PFOS, PFOA, PFNA, PFDA, PFHxS, PFHPS), even though only PFHPS, PFDA, and PFNA were statistically significantly different at 0.05 level. While we are unsure why this is the case, it may be that Wisconsin has less exposure to PFAS in the environment and/or in products used by its residents. PFOS was among the most prevalent of the PFAS compounds measured among the SHOW sample and had the highest geometric mean and percentile concentrations. This was also true among NHANES, as was seen among nearly all prior biomonitoring studies, including those who tested high-risk populations such as firefighters and communities near contamination sites [24-26,46,47]. Higher serum concentrations levels of PFOS are not surprising due its (and PFOA’s) U.S. production since the 1940s, which peaked between 1970 and 2002. PFOS and PFOA were the most widely used PFAS compounds in consumer products up until recently [10].

Males and older age were associated with higher levels of PFAS in SHOW across all PFAS compounds analyzed, with few exceptions. This trend held true in NHANES, and in other studies. For example, communities tested near military bases in Pennsylvania [45] found PFOS, PFOA, and PFHxS were all higher in males, and all increased with age. Those tested living nearby a PFAS manufacturing facility in New Jersey saw similar increases in PFAS concentration with age and were higher among males [24]. Olsen et al. similarly found sex and age trends as seen in NHANES [48]. Lower concentrations in females may be explained by greater excretion rate through menstrual blood loss and lactation [49]. We would expect PFAS concentrations to be higher among older adults compared to younger adults due to the bioaccumulation that occurs as one ages, in conjunction with potentially higher exposure >10 years ago before PFAS started being manufactured oversees and phased out of products.

Both SHOW and NHANES saw similar trends in PFAS concentrations by race, with non-Hispanic whites having higher geometrics means compared to non-whites. While in SHOW, this trend remained across percentiles, the NHANES sample saw opposite trends at the 90^th^ and 95^th^ percentile for some compounds (PFHPS, PFDA, PFNA, PFOS), where non-whites had higher PFAS concentrations than non-Hispanic whites. This difference may be due to the small non-white sample in SHOW and therefore an inability to adequately capture potential racial differences in exposure and risk. Non-whites only account for 18.7% of the SHOW sample, compared with approximately 35% in each NHANES sample. While the SHOW study oversampled non-whites in 2019, they did not conduct oversampling of non-whites during their statewide sampling years (2014-2016) used in this study. Due to Wisconsin’s proportionately small non-white population, the non-white sample in the SHOW study sample is small. This likely resulted in little variability within the non-white subpopulation which may have resulted in an unrepresentative substratum of non-whites in the state. As such, those who were included are weighted to represent a larger proportion of their racial/ethnic subpopulation. Racial and ethnic minorities, and those of lower socio-economic and education attainment, are more likely to reside near industrial and contamination sites that are at greater risk of exposure to environmental contaminants of concern, such as PFAS. This was seen in biomonitoring of a high-risk community due to manufacturing in Alabama where PFOS levels were ∼2 times higher in African Americans compared to whites in the study [26]. We would expect a similar trend in SHOW, but additional testing or sampling is needed.

The SHOW’s relatively small substratum of non-whites was a limitation of the study. This may have resulted in a less balanced comparison to NHANES and limited the study’s ability to identify racial disparities relating to PFAS that may exist in Wisconsin. While race may be an important predictor of PFAS exposure, there are other demographic differences between SHOW and NHANES that may have contributed to differences in the comparison of results. On average, SHOW participants had a slightly higher education level and income to poverty ratio than NHANES participants in all sampling years. It is well-known that lower income status is associated with increased adverse environmental exposures [50], which may have contributed to the lower levels of PFAS in SHOW participants than may exist in the state’s population. There is evidence that higher PFAS exposure is associated with lower educational attainment [46].

While the SHOW 2014-2016 sample is representative of the state, its primary sampling unit was at the county level, and only 9 counties were represented. Selection criteria used for SHOW’s sampling frame to ensure representation was based on socioeconomic status and population density; geographical representation was not a factor. This is a significant limitation of the study as northern regions of the state, and more isolated areas, are not represented. Future PFAS biomonitoring in the state should include oversampling of non-whites and geographical representation to adequately capture varied land use throughout the state; ensuring unique areas in central sands regions and northeast areas with karst geology are included, due to potentially vulnerable groundwater contamination. Additionally, data on education and smoking status were unavailable for 18- and 19-year-olds for NHANES, so some demographic comparisons do not include these individuals. Lastly, this study was cross-sectional, with serum concentrations from over 6 years ago. Hence, not only can causality not be inferred, but changes over time are not captured, and current PFAS concentrations among the state’s population today may be different than seen in this study. Future PFAS biomonitoring in Wisconsin should consider repeat testing, especially as short chain PFAS become more widely used and long chain PFAS are phased out. While this study has several limitations, it has many strengths. This is the first statewide representative cohort for which we have baseline PFAS serum concentrations. Studies-to-date have conducted biomonitoring on specific cohorts (i.e. California Teachers Study), among high-risk occupational workers (Firefighters), or communities residing near a contamination site.

While Yu et al. conducted statewide biomonitoring, they relied convenience testing from clinical and lab sera [44], whereas SHOW relied on probability sampling to produce a statewide representative sample with weights. This study also tested a wider range of PFAS compounds with LLOD for many. This was an important contribution to the field and increased our understanding of the extent other PFAS compounds are in the environment and in our bodies. These data suggest that Wisconsin residents may not be disproportionately burdened by PFAS contamination compared to the wider US population. More research is needed to determine the extent of PFAS exposure in Wisconsin which ensures geographical variation and adequate oversampling of non-whites. This is the beginning of our understanding of PFAS exposure among Wisconsin residents, but importantly, it offers a baseline prevalence of PFAS. Future directions include utilizing the rich SHOW survey data to better characterize PFAS serum levels based on diet, housing, and other factors. These findings can help state agencies in resource allocation for additional PFAS biomonitoring, and direct resources where most needed.

Additionally, residential address and residential history is known for SHOW participants, which increases the capacity to study cumulative exposure to PFAS through neighborhood-level contextual factors like industrial sites and nearby land uses through the adult life course. Finally, longitudinal follow-up should be conducted in the future to track changes in PFAS burden in the population over time.

## Supporting information

(see Supp. Tables 1-5)

(see Supp. Tables 1-5)

(see Supp. Tables 1-5)

(see Supp. Tables 1-5)

(see Supp. Tables 1-5)

## Data Availability

The datasets generated during and analyzed during the current study are not publicly available due to HIPAA protections for SHOW participants but may be available from the corresponding author on reasonable request with IRB approval.

## Acknowledgements

The authors would like to thank all the SHOW investigators who made this work possible, WSLH staff who contributed to the creation of this data, and the SHOW participants.

## Author Contributions

Conceptualization: A.S., N.S., K.M.; Methodology: N.S., M.L., A.S., Sample Analysis: N.S., M.L., Data Analysis: A.S., R.P.; Draft Preparation: A.S., R.P., N.S., K.M.; Review and Editing: A.S., R.P., R.I., J.M., K.M.; Supervision: K.M., N.S.; Funding Acquisition: A.S., N.S.

## Ethical Approval

The SHOW protocol and informed consent documents are approved by the Health Sciences Institutional Review Board of the University of Wisconsin-Madison. Participants in SHOW gave consent to their information being used for research prior to this study.

